# Exploring a targeted approach for public health capacity restrictions during COVID-19 using a new computational model

**DOI:** 10.1101/2022.11.28.22282818

**Authors:** Ashley N. Micuda, Mark R. Anderson, Irina Babayan, Erin Bolger, Logan Cantin, Gillian Groth, Ry Pressman-Cyna, Charlotte Z. Reed, Noah J. Rowe, Mehdi Shafiee, Benjamin Tam, Marie C. Vidal, Tianai Ye, Ryan D. Martin

## Abstract

This work introduces the Queen’s University Agent-Based Outbreak Outcome Model (QUABOOM), a new, data-driven, agent-based Monte Carlo simulation for modelling epidemics and informing public health policy in a wide range of population sizes. We demonstrate how the model can be used to quantitatively inform capacity restrictions for COVID-19 to reduce their impact on small businesses by showing that public health measures should target few locations where many individuals interact rather than many locations where few individuals interact. We introduce a new method for the calculation of the basic reproduction rate that can be applied to low statistics data such as small outbreaks. A novel parameter to quantify the number of interactions in the simulations is introduced which allows our agent-based model to be run using small population sizes and interpreted for larger populations, thereby improving computational efficiency.

## 1. Introduction

The COVID-19 pandemic has globally impacted society economically and socially. Since emerging in 2019 in Wuhan, China (Wu and McGoogan, 2020), the novel coronavirus SARS-CoV-2 has spread worldwide and led to the implementation of non-pharmaceutical policy interventions by public health authorities including social distancing, lockdowns, quarantine, and mask mandates.

In March 2020, Canada declared a COVID-19 pandemic (Statistics Canada, 2022a) which led to policy mandates being implemented, removed and re-implemented as the pandemic evolved over time. In Canada, all industrial sectors were negatively impacted by the pandemic. Particularly a large portion of small businesses (fewer than 99 employees) reported a 40% or more decline in revenue (Tam and Johnston, 2020). Small businesses comprise 98% of employer businesses in Canada (Sood et al., 2021), highlighting the importance of understanding how they affect the development of epidemic outbreaks.

Mathematical modeling has become an important tool in policy making for infectious disease control to mitigate and suppress the health impacts of COVID-19 (Ferguson et al., 2020; van der Vegt et al., 2022; Ndaïrou et al., 2020). An abundance of epidemic modelling techniques exist, with compartmental-based models, such as susceptible-infected-recovered (SIR), being the most commonly used in epidemiology (Weissman et al., 2020; Tolles and Luong, 2020). Modelling techniques are often implemented using ordinary differential equations (He et al., 2020; Chowell et al., 2004), networks (Newman, 2019), and agent-based models (Ferguson et al., 2020). Our work presents a Monte Carlo agent-based model that includes network-like constraints between its agents in order to reduce the full random-mixing assumption implicit in differential equation based models and results in more realistic social mixing.

While COVID-19 has been at the forefront of epidemiology research, very little modeling has been conducted focusing on small-scale populations. Our technique was developed with the capability of modeling a city the size of Kingston, Ontario, Canada, with a population size of approximately 130 000. The city also includes a relatively large student population (approximately 20 000), also incorporated in the model. This first publication presents, to our knowledge, a new model which focuses on examining how capacity restrictions on businesses of different sizes affect an epidemic outbreak. We present a method to inform public health capacity restriction decisions and examine how these change as a function of the vaccinated fraction of the population.

In Section 2, we describe the general software framework developed to model the spread of COVID-19. In Section 3, we describe how we use the code to determine a prescription for setting capacity restrictions. In Sections 4 and 5, we then present and discuss results of simulating various capacity restriction scenarios.

## 2. Agent-based modelling software framework

In our model, agents represent individuals that interact and can spread an infection. Agents are given a set of properties, such as age, that will affect their probability of getting infected and the outcome of any infection.

The model is referred to as the Queen’s University Agent-Based Outbreak Outcome Model (QUABOOM), and is implemented using object-oriented code written in Python 3.8, which will be made available as open source code. The simulation tracks a population of agents during an epidemic. An initial number of agents, *N*_0_, are infected at the beginning of the simulation. Each day within the simulation, agents are given opportunities to interact with each other in different “interaction sites” which represent establishments such as households, work places, restaurants, and grocery stores. Interactions between agents are modelled with a Monte Carlo simulation, through probabilities of attending an interaction site and transmitting the virus. Networks of agents are formed by associating subsets of agents to specific interaction sites. Various public health policies, such as masking, testing, quarantining, and lockdowns can also be implemented in the simulation.

The hierarchy of classes developed for the model are shown in Fig.1. The Simulation class creates and calls other classes to manage the agents, their interactions, and any public health policy in place. The agents are created by the Population class as Person objects. Interactions between agents are facilitated by the InteractionSites class. The InteractionSites class models how agents interact with each other in different interaction sites and is how agents spread the virus. The Policy class is used to implement public health policy. The following sections describe the details of the various classes implemented in the code.

**Fig. 1:**
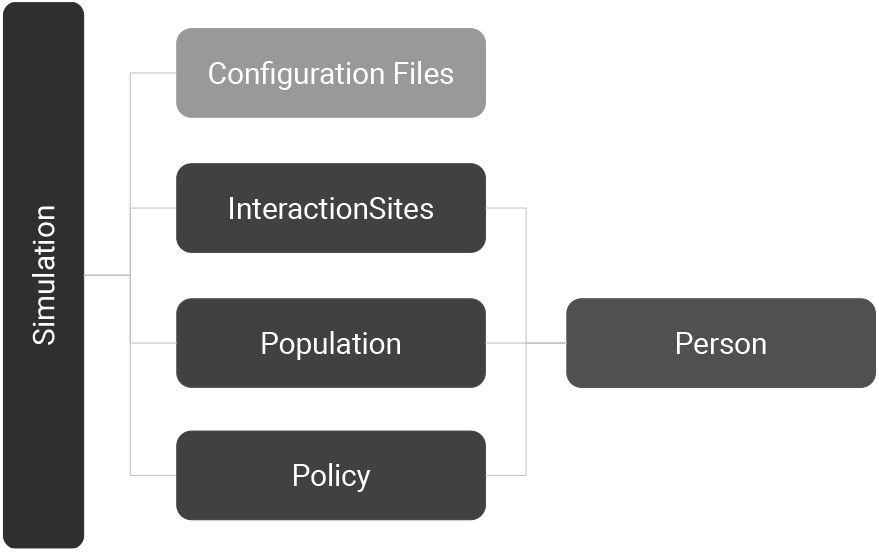
Software framework used for mathematically modelling COVID-19 with QUABOOM, in Kingston, Ontario, Canada. Each dark box corresponds to a class in the code which can be configured through a set of configuration files in TOML format.

### 2.1. Simulation class and configuration files

The Simulation class initializes all relevant classes and organizes the interaction between agents from the Person class as they interact in the various interaction sites. The Simulation class tracks daily cumulative counts from the simulation for the following quantities:

- Susceptible agents
- Infected agents
- Recovered agents
- Dead agents
- Hospitalized agents
- Infected student agents
- Agents tested
- Agents in quarantine
- Agents waiting to get tested
- Daily tested agents
- Daily quarantined agents
- Daily infected agents

The Simulation class uses parameters specified in two configuration files and distribution data from properties of the Kingston population. The first configuration file controls simulation-wide parameters such as the size of the population, the number of initially infected, any public health policies to implement, and the distribution of the populations’ age. The second configuration file controls the disease parameters, including symptom severity distributions, length of stay in hospital/ICU, and information regarding transmission probabilities. The disease parameters allow for multiple variants to be defined by treating each variant as a new virus type. This gives the simulation the ability to be used for epidemiological purposes further than COVID-19 by modelling infectious virus epidemics with different properties than COVID-19.

### 2.2. Person class

The Person class represents the base agent object within the simulation. Each agent has a set of unique attributes, set by the Population class to reproduce distributions from the configuration files. The interactions, managed by the InteractionSites class, that an agent has with other agents are stored in that agent’s attributes, allowing for contact tracing to be simulated. Each agent also has a “compliance” parameter, used to account for individuals’ varying adherence to public health policies. This can change during the simulation but is set to be constant for simplicity within the scope of this paper. An agent’s initial compliance is determined by different factors, including risk due to pre-existing conditions and age. Compliance can also be updated during the simulation to reflect real world tendencies during lockdowns and lifting of restrictions.

### 2.3. Population class

The Population class initializes the agents, which are of type “Person” from the Person class in order to reproduce a given distribution of ages in the population. The Population class also sets the initial state of agents (infected, susceptible, vaccinated), and is then responsible for tracking the state of all of the agents in the population throughout the simulation. The Population class also places agents into households. The Simulation class can then query the Population class at each time step of the simulation to track the current number of agents in the various states.

### 2.4. InteractionSites class

Interactions between agents are modelled as taking place at various interaction sites and are managed by the InteractionSites class. There are three main interaction site levels that are designed to model different types of sites. “Level A” represents facilities, such as clothing stores and restaurants, for which a given individual might have several locations with which they are associated. “Level B” corresponds to sites for which an individual may have one or two instances with which they are associated, such as grocery stores, gyms, and gas stations. Finally, “Level C” corresponds to sites where an individual associates singularly, such as a workplace or school. In addition, the code allows smaller scale dynamics to be studied by permitting for a population of students to be included in the general population. When the student population is enabled, additional types of interaction sites to model lecture theaters, study areas, food areas, student residences, and student housing are included in the simulation.

In the simulation configuration file, the following parameters for each level of interaction site can be estimated:

- The number of interaction sites, *n*, of a particular level. This also determines the number of agents associated with each interaction site of a given level, as the agents in the populations are evenly distributed over the sites.
- The “attendance” probability, *p*, is the probability that an agent will go to that level of interaction site on a given day. For example, for Level B sites, we use a mean of *p*_*B*_ = 2/7, corresponding to attending a grocery store type of establishment twice per week, assuming the same probability of attending a site on any given day, ignoring any weekend effects.
- The “loyalty” mean and standard deviation are used to draw a normally distributed random number for the number of sites of a given level with which an agent is associated.

Agents are initialized to be associated with specific interaction sites by the InteractionSites class. The number of specific sites of a particular level that an individual can attend is fixed and drawn from a normal distribution whose parameters (loyalty mean and standard deviation) are specified in the simulation configuration file. The same distribution is used for all agents, independent of age, and it is assumed that the over-estimations and under-estimations of attending a specific site even out. An agent will then repeatedly “attend” the same set of interaction sites with a probability, *p*, of going each day. Different functions manage interactions between agents going to the same interaction site and interactions between members in the same household. This feature aims to replicate real world behaviours, where an individual is more likely to visit the same public locations, for example a workplace, and interact with the same individuals, for example at a house.

For each time step of the simulation, a list of agents that will interact in each site is built based on the attendance probability, *p*, for that level of site. For each agent, a number of interactions is drawn from a triangular distribution peaked at zero and that decreases linearly to the maximum number of contacts an agent could have in that site. The maximum number of contacts is the total number of agents that will visit the site that day divided by hours, *h*, to account for the fact that the agent can only interact with those that are at the site at the same time (assumed to be one out of *h* hours the site is open). We assume that most sites are open for 12 h and customers attend for one hour on average, so that *h* = 12 h^1^.

In a population of *N* agents, where there are *n*_*B*_ interaction sites of level B, and agents have a probability to attend a site per unit day, *p*_*B*_, one out of the *h*_*B*_ hours that site is open each day, the average number of interactions per hour per interaction site, *i*_*sh,B*_, is given by:

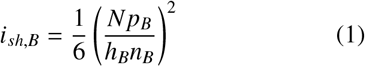

where the agents are distributed such that each site has the same number of agents associated with it. The factor of 1/6 accounts for the mean of a triangular distribution being 1/3 of its maximal value and the number of interactions are double counted if they are not divided by the number of agents in an interaction, 2. Equation 1 assumes each site has, on average, the same number of agents.

Individual contact networks are naturally generated between agents that visit the same interaction sites and are maintained throughout the simulation. This feature allows the model to naturally incorporate network-like features among the agents.

When an interaction occurs between two agents, there is a probability of transmitting the virus, *P*_*T*_, if one agent is in the infected state and the other is in the susceptible state. We model *P*_*T*_ as a fixed number that should be a direct property of the virus. We examine the results of our simulation as a function of *P*_*T*_, since this probability will necessarily have a range of values depending on the variant and the two agents in the interaction.

Furthermore, the chance of the virus spreading during an interaction is dependant on the vaccination status of both agents and the corresponding vaccine-dependent efficiencies (Pfizer-BioNTech Comirnaty, Moderna Spikevax, Oxford-Astrazeneca Vaxzevria). In addition, we consider whether one or both agents are wearing a mask and assign different reduced infection probabilities to wearing different efficiency masks (surgical or cloth). These additional efficiencies (vaccination and masking) affect the effective probability of transmission.

### 2.5. Policy class

The Policy class is responsible for managing the following public health policies in the simulation:

- Masking mandate: whether or not agents should be wearing masks (affecting spread probability).
- Lockdown: in a lockdown, only houses and level B sites are open, such as grocery stores (affecting number of interactions).
- Testing: when testing is turned on, symptomatic agents must get tested and quarantined if positive.
- Students: whether or not agents with the ‘student’ profession will be involved in the simulation.

Each policy can be triggered on or off by a daily case count, a specific date, or set to be static by initializing the mandate at the beginning of the simulation. Individual policy adherence is also affected by a person’s compliance score.

A model for COVID-19 testing is implemented each day by selecting a group from the population wait-listed to get tested due to apparent infection symptoms and informing them if they are infected with COVID-19. There is a random chance that individuals will go for testing without COVID-19, to simulate those with symptoms from other infections. Individuals who test positive are quarantined, which restricts them from engaging in interactions with other individuals for the set quarantine time. Currently the model does not account for inaccuracies of the tests themselves, although it has no effect on the results presented herein. Similar to tests, a set number of vaccinations can be administered each day or set at the beginning of the simulation. Vaccines largely reduce the transmission probability for both individuals in an interaction. The simulation can implement multiple vaccines with varying efficacy in order to study the effect of varying or waning efficacy.

### 2.6. Baseline parameters and example output

Our default configuration files specify a number of “baseline” parameters for the simulation and the virus properties. The most relevant disease parameters are shown in Table 1 and correspond to literature values for the original strain of SARS-CoV-2, while the most relevant simulation parameters are illustrated in Table 2.

**Table 1:** Disease baseline parameters, based on original strain of SARS-Cov-2. Onset of symptoms is the number of days before symptoms (Li et al., 2020); mild days is the number of days of symptoms onset for a mild case(Li et al., 2020), hospital days is the onset days to acute respiratory distress syndrome (ARDS) (Huang et al., 2020) plus days to onset of symptoms; ICU days is the days to symptom onset plus time to hospital admission for survivors plus length of ICU stay for survivors (Zhou et al., 2020); die days is the time to onset plus time from illness onset to death for non-survivors (Zhou et al., 2020); and the incubation period of COVID-19 is the days between exposure and symptom onset (Statistics Canada, 2022b).

| Disease parameters | Value |
| --- | --- |
| Transmission probability | 0.35 |
| Symptoms onset | 4 - 7 days |
| Mild days | 4 - 7 days |
| Hospital days | 12 - 21 days |
| ICU days | 16 - 29 days |
| Die days | 16 - 29 days |
| Incubation | 1 - 14 days |

**Table 2:** Main baseline simulation parameters and default policies. Where the default probability of testing is the percentage of the population that will get tested if they have COVID-19, s_A_ is the default amount of agents associated with one site A, p_A_ is the probability an agent will attend an interaction site A each day, and l_A_ is how likely an agent is to visit the same interaction site (mean) or visit various interaction sites (std.); each type of interaction site has its own average size, attendance probability, and loyalty (mean and std.).

| Agent parameters | Value |
| --- | --- |
| $N$, Population size | 10 000 |
| Initial infected | 10 |
| Initial vaccinated | 0 |
| Initial mask mandate | True |
| Initial lockdown mandate | True |
| Initial testing mandate | True |
| Initial student mandate | False |
| Quarantine time | 14 |
| Default probability of testing | 100% |
| $s_A$, A site size | 400 |
| $p_A$, A attendance probability | 1/7 |
| $l_A$, A Loyalty (mean and std.) | $10 \pm 2$ |
| $s_B$, B site size | 1000 |
| $p_B$, B attendance probability | 2/7 |
| $l_B$, B Loyalty (mean and std.) | $1 \pm 1$ |
| $s_C$, C site size | 170 |
| $p_C$, C attendance probability | 5/7 |
| $l_C$, C Loyalty (mean and std.) | $1 \pm 0.2$ |

Fig. 2 shows a sample of outputs from the first 50 days of a simulation run with the baseline parameters. The number of agents in each state is shown as a function of days in the simulation. As more agents become infected over time and there is a smaller susceptible population, less agents can become infected and the epidemic comes to an end, as expected. Five different simulations were performed and the output of each were overlayed. The resulting curves were then averaged to minimize statistical fluctuations in any given simulation and were estimated to a 95% confidence interval.

**Fig. 2:**
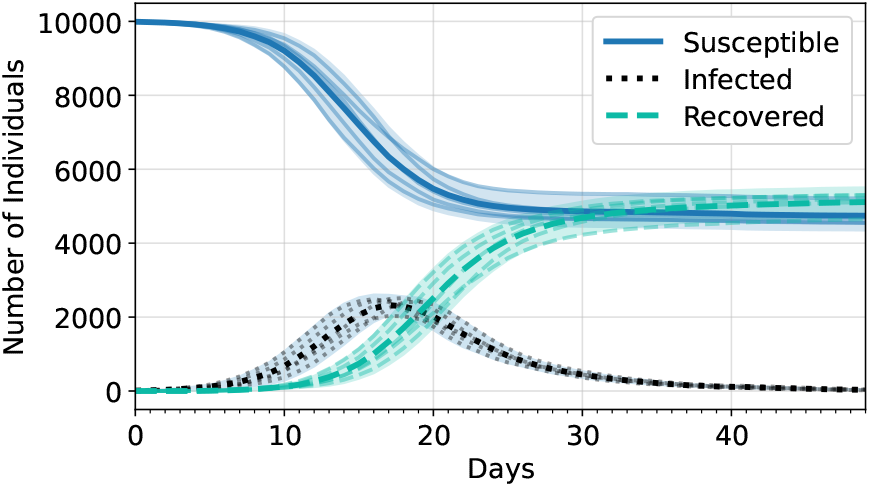
Output of epidemic with a population size of 10 000, a masking probability of 0.8, an attendance probability of 2/7 and a probability of transmission of 0.25. Each compartment (Susceptible, Infected, and Recovered) has a superposition of 5 runs (faded lines), the average of all the runs (in a thick line), and a 95% confidence interval (in the shaded region).

Fig. 3 highlights the code’s ability to simulate lockdowns, which restricts agents to attend only level B interaction sites when a certain percentage of population becomes infected and then turns off when there are less active infections. This showcases how lifting capacity restrictions too early can result in another large wave of infections, thus requiring another lockdown to reduce the number of active cases.

**Fig. 3:**
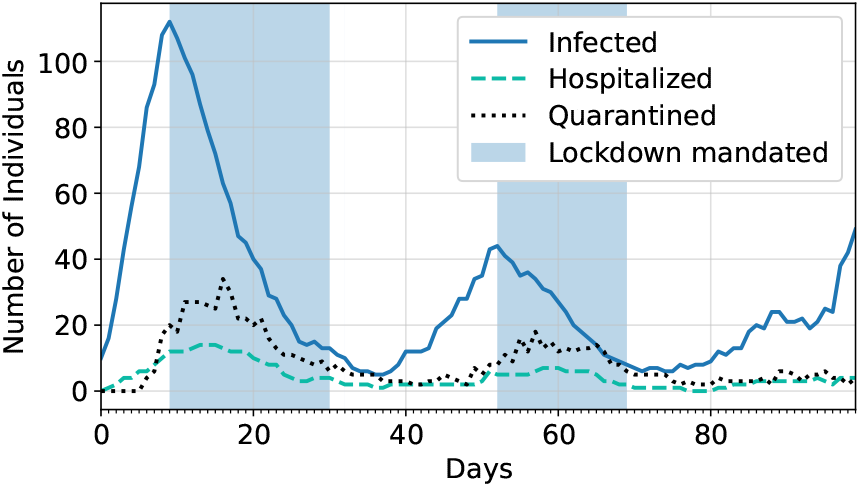
Sample of a simulation with masking and testing always mandated, where the lockdown is triggered when 1% of the current population is showing infection symptoms and is turned off when below 0.1%.

Fig. 4 demonstrates how a variant with a different probability of transmission can be introduced to the simulation and can quickly dominate by becoming the new main infection. A limitation of the current model is its inability to allow re-infection, including infections from different variants. Since this work focuses on the first large peak of infections, it does not impact the results, but future work includes the addition of re-infection to mitigate this.

**Fig. 4:**
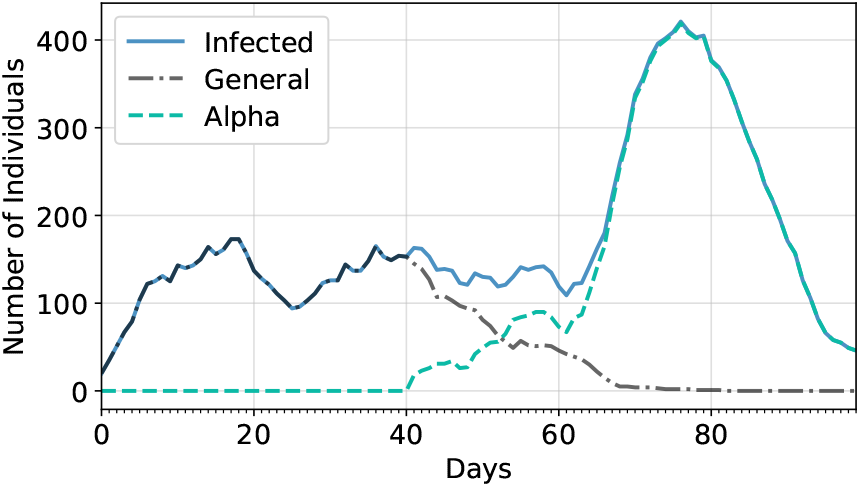
Sample of a simulation with a lockdown, masking and testing always mandated, where a more infectious variant, called alpha, is introduced on day 40. The alpha variant is set to be 1.8 more contagious than the original variant.

## 3. Methodology for understanding capacity restrictions

### 3.1. Overview

In this section we describe how the model can be used to understand the effect of capacity restrictions during a lockdown. In particular, we focus on understanding how to determine capacity restrictions that would prevent a small number of infections from resulting in a large epidemic outbreak across the population. The model is configured to approximate the conditions in Kingston at the beginning of the COVID-19 pandemic when only essential businesses were open.

A lockdown mode is implemented such that only one type of interaction site (level B, grocery store-like) is available for agents to infect each other outside of their households. In the following sub-sections we describe how we define a basic reproductive number for the simulation, which is then used to define an epidemic threshold based on the number of agents infected at the peak of an epidemic.

Our results focus on understanding how this epidemic threshold varies as a function of two fundamentally independent parameters of the simulation: the probability of transmission, *P*_*T*_ (a property of the virus in a given environment) and the average number of interactions that agents have per unit time per interaction site of a given level, *i*_*sh,B*_ (a quantity that capacity restrictions change). By considering the epidemic threshold as a function of *i*_*sh,B*_ (see equation 1), the results of a simulation can be interpreted for different population sizes, *N*, different probabilities of attending an interaction site per day, *p*_*B*_, and different number of interaction sites, *n*_*B*_. This is a result of distributing agents equally among the interaction sites and having only one level of interaction site open. This setup allows us to explore a rich parameter space by only varying *i*_*sh,B*_ and *P*_*T*_, while using a relatively small number of agents, *N* = 10 000, to increase computational efficiency.

For each set of input parameters (*i*_*sh,B*_, *P*_*T*_), we run five simulations (as in Fig. 2). Each simulation outputs a number of metrics, including susceptible, infected, quarantined, and recovered agents each day. These metrics are then averaged over the five simulations and used to compute additional parameters, such as the basic reproduction rate and average epidemic curves.

### 3.2. Definition of ℛ_0_

In order to define an epidemic threshold (section 3.3), based on the number of infected cases in a simulation, we introduce an effective ℛ_0_ (basic reproductive number) that can be computed from both the simulation and real data for small epidemics and populations. In epidemiology, the basic reproductive number is defined as the average number of secondary infections by one infected individual in a fully susceptible population (Hethcote, 2000). Our definition is based on a standard differential equation 3-compartment model for the number of susceptible (*S*), infected (*I*), and recovered (*R*) people in a population of *N* individuals (Hethcote, 2000):

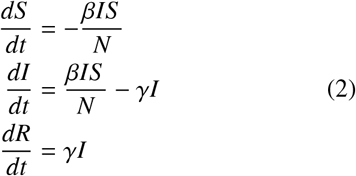

where *β* is the infection rate (per unit time) and *γ* is the recovery rate (per unit time). A reproductive number can be defined as:

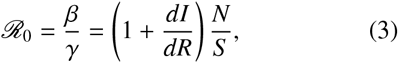

such that the number of infected agents will grow with time if ℛ_0_ > 1. In other words, to first order, if the rate at which people are being infected is larger that the rate at which they recover, the number of infected people will grow.

The change in the number of infected and recovered cases (*dI* and *dR*) can be obtained from either the simulation or real population data. We explicitly assume that ℛ_0_, which depends on the ratio of *dI* to *dR*, is unaffected by under-counting cases in the real data. We also take *N*/*S* ∼ 1, as we consider data only at the beginning of the pandemic.

The values of *dI* (change in infected/active cases) and *dR* (change in recovered cases) are taken at different instances in time in the simulation (or the data) so that the change in recovered individuals measured on a given day corresponds to the same day as those individuals were infected. We implement a lag, *L*, as the number of days between the measures of *dI* and *dR*. We treat *L* as an unknown parameter that depends on various delays in reporting active and resolved cases in the real data. In the simulation data, the value of *L* is expected to be close to the average time to recover from an infection.

In practice, we apply a running average with a window, *w* = 7 days, to the values of *dI* and *dR* before computing ℛ_0_. Due to statistical fluctuations in small epidemics, it is possible for the value of ℛ_0_ to be negative. We thus define our value of ℛ_0_ as the mean of the non-negative values computed over a given range of days, typically of order 2 months. The lag is then varied until a minimum in the standard deviation of the ℛ_0_ values is found. We found that our simple definition produced a stable and consistent value of ℛ_0_ in the small epidemics that we have studied, as illustrated in Fig. 5 with data from Kingston, Ontario.

**Fig. 5:**
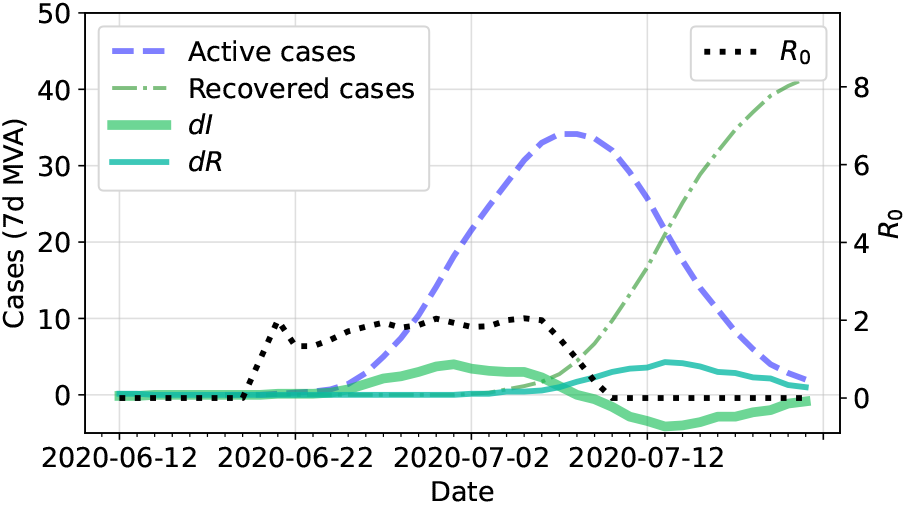
Epidemic data for Kingston, Ontario, from July 2020 that were used to calculate a value of R_0_ (right axis, dotted black line). Data from Ontario Public Health (2022) were obtained for active/infected (left axis, dashed line) and resolved cases (left axis, dashdot line). From those, the changes dI and dR are computed (left axis, solid lines), and a lag of approximately 12 days can be observed between those values.

Using this methodology, we infer a value of ℛ_0_ = 1.7 ± 0.5, as measured using 40 days of data from Kingston centred around an outbreak that occurred in July 2020, as illustrated in Fig. 5. This corresponds to the mean and standard deviation of the non-negative ℛ_0_ values calculated with a lag of *L* = 12 days between the change in active cases and the change in resolved cases.

### 3.3. Defining epidemic threshold in the simulations

In order to examine factors that can trigger a large epidemic outbreak, we define an epidemic threshold based on the number of agents infected at the peak of the epidemic. We determine that threshold value using ℛ_0_. We verified that our definition of ℛ_0_ behaves as expected and leads to large epidemics when ℛ_0_ > 1, as illustrated in Fig. 6.

**Fig. 6:**
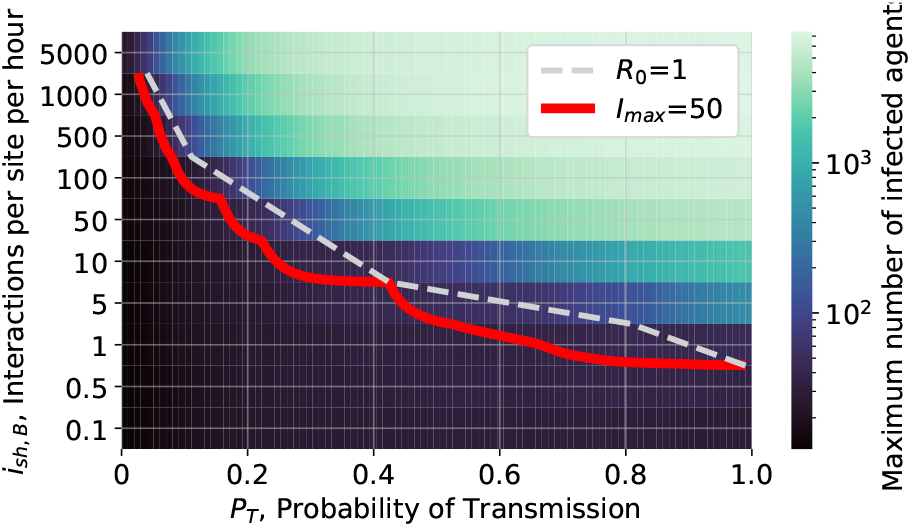
A two-dimensional plot of mean max infected, I_max_ (logarithmic colour scale), as a function of the probability of transmission, P_T_, and the average number of interactions in level B sites per hour per site, i_sh,B_, (logarithmic) in a population of N = 10 000. The dashed white line corresponds to defining an epidemic threshold based on ℛ_0_ >= 1. The solid red line shows that an equivalent and smoother threshold can be defined with the value I_max_ = 50 in this particular configuration of simulations.

In practice, the value of ℛ_0_ is subject to statistical fluctuations that are due to the small values of *dR* and *dI* for small epidemics, especially near the threshold value. By defining an epidemic threshold based on the maximum number of agents infected in a population, we compute the number of agents infected at the peak of an epidemic (averaged over five simulations) and use this “max infected”, *I*_max_, as a measure of the size of the epidemic outbreak for a given set of simulation parameters. We then introduce a threshold, *T*_*I*_, for the largest epidemic that we are willing to tolerate. The *T*_*I*_ value is chosen from *I*_max_ as seen in Fig. 6, allowing for a qualitative threshold to be chosen which is below ℛ_0_ = 1.0 (where we expect the number of infections to grow). In this work, a baseline value of, *T*_*I*_ = 50 is used, corresponding to 0.5 % of the simulated population being infected and a value of ℛ_0_ = 1.0, as illustrated in Fig. 6.

## 4. Results: Behaviour of the epidemic threshold as a function of model parameters

Fig. 6 shows a two-dimensional plot of the maximum number of agents infected in an epidemic, *I*_max_, averaged over 5 simulations (logarithmic colour scale) as a function of the probability of transmission, *P*_*T*_, and the average number of interactions in level B sites per hour per site, *i*_*sh,B*_. The data in Fig. 6 were smoothed by a Gaussian filter (Virtanen et al., 2020) applied to each row. The smoothed data were then used to determine a contour (in solid red) at the value *T*_*I*_ = 50. The dashed grey contour line was determined from the same set of simulations using the condition that ℛ_0_ = 1 and used to determine the threshold value *T*_*I*_, which can be calculated more reliably than ℛ_0_.

The results in Fig. 6 make intuitive sense; when *P*_*T*_ and *i*_*sh,B*_ are high, a large fraction of the population is infected at the peak of the infection. Conversely, if *P*_*T*_ and *i*_*sh,B*_ are are low, there are no large outbreaks. The contour line then gives the number of interactions per site per hour that one can tolerate for a given probability of transmission. We discuss in the next section how these results can provide guidance to public health units.

Fig. 7 shows how the contour determined in Fig. 6 changes as different fractions of the populations are vaccinated at the beginning of the simulation. Three vaccine efficacies of 94.1 %, 91.3 %, and 76 % were used to simulate the population being fully vaccinated; where 35 % of the population received Moderna, 60 % Pfizer and 5 % Astrazeneca, with the respective efficacies. Since the simulation is run over a short time period, the vaccine efficiencies are assumed to be constant and do not account for varying efficiency for different variants. As expected, more interactions can be tolerated as the population is vaccinated and this information could be used to inform the lifting of capacity restrictions.

**Fig. 7:**
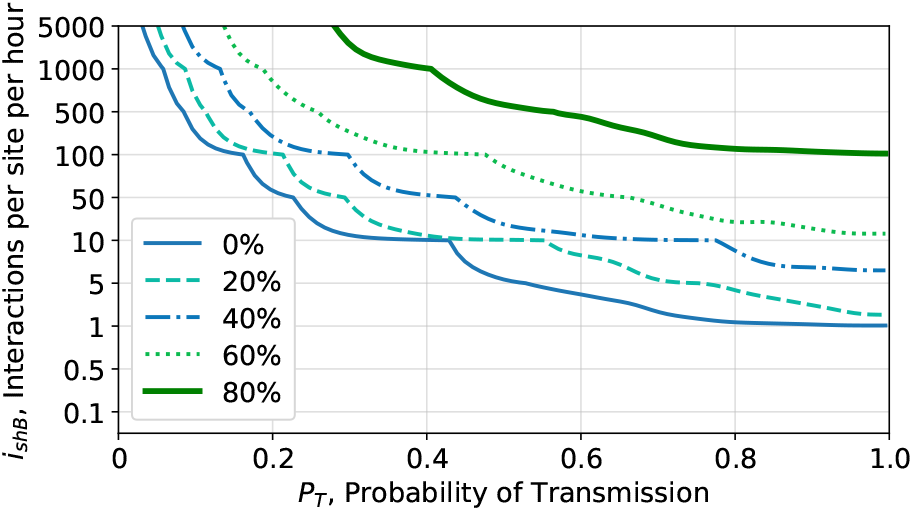
Epidemic thresholds as different fractions of the population are vaccinated as a function of probability of transmission, P_T_, and number of interactions per site per hour, i_sh,B_.

## 5. Discussion: Considerations in determining capacity restrictions

While the simulations were run with a population of 10 000 agents, we can interpret the contour on a population the size of Kingston, with a population 10 times larger. Consider a case when the probability of transmission is *P*_*T*_ = 0.2 so that the epidemic threshold will be crossed if the value of *i*_*sh,B*_ ≳ 50 interactions/site/hour, when referring to Fig. 6. In this case, a public health unit may decide that if people go once per week to a level B interaction site (e.g. a grocery store), *p*_*B*_ = 0.14, for one hour out of the *h*_*B*_ = 12 h that they are open, then, the largest number of sites that should open is given by:

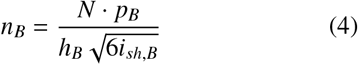

resulting in *n*_*B*_ = 69 sites that could open for a population of *N* = 100 000. This is a sizeable number of sites open, where each would have an average of *N*/*n*_*B*_ = 1455 customers associated with that site. One can then refer to Fig. 7 to understand how restrictions can be lifted as the population becomes vaccinated.

A public health unit may instead decide to open sites based on the assumption that people go twice per week, *p*_*B*_ = 0.28, in which case they could open half as many sites, *n*_*B*_ = 35, each with half as many customers associated (727). Similarly, a public health unit in a town of 10 000 could use the same simulated data to decide to open 10 times more sites (or that they do not need any significant capacity restrictions).

This model and the interpretation of its outputs in terms of the mean number of interactions per hour per interaction site can provide useful guidance to public health units to create tailored measures. For example, rather than implementing large scale lockdowns across diverse establishments (different level interaction sites) one should tailor capacity restrictions to those businesses that have the largest impact. In our model, when the probability of transmission is 20%, the small interaction sites, with few customers associated with each, lead to low values of *i*_*sh,B*_. Small businesses (small stores, etc) as interaction sites do not contribute significantly to an epidemic outbreak through the interaction between their customers.

Conversely, we find that it is large interaction sites, with many interacting agents, that lead to large epidemic outbreaks. This would suggest that shutting down businesses with few employees while leaving businesses with many employees open, may not have been the optimal approach for implementing restrictions, from an epidemic outbreak consideration. Our study suggests it could be more effective to shut down the large businesses and encourage the population to support small businesses while they are otherwise working remotely.

When we modify the simulation presented in Fig. 6 to include additional interaction sites of level A (*n*_*A*_ = 250, *h*_*A*_ = 12, *p*_*A*_ = 2*/*7), each with 400 agents associated, the contour that is inferred is effectively unchanged, since this corresponds to adding approximately (only) *i*_*sh,A*_ = 15 interactions*/*site*/*hour. This supports the conclusion that one can open a relatively large number of sites that have of order a few hundred agents associated with them, as long as the probability of transmission between two agents in those sites is relatively small, as it would be in most small businesses.

Consider the decision to open a few “essential” large workplaces with a high transmission probability, such as a warehouse, in a population with 100 000 agents. We can model agents as going to a site for the entire time that it is open, *h*_*B*_ = 1, five out of seven days per week, *p*_*B*_ = 5*/*7, and have a high transmission probability *P*_*T*_ = 0.5 to model a workplace. In this case, one finds that the number of interaction sites must be about *n*_*B*_ ∼ 30 000, have *i*_*sh,B*_ ≈ 1 and be on the contour in Fig 6. This does not mean that one can open 30 000 ware-houses; rather, it means that one can only open interaction sites where the average number of agents associated with any site is given by *N/n*_*B*_ = 3.4. In other words, interaction sites that results in large transmission probabilities and high attendance need significantly stronger restrictions in order to prevent an epidemic. If one only considered such workplaces, then one could conclude that wide-ranging lockdowns are required. In Ontario, approximately 50% of businesses have less than four employees (Statistics Canada, 2020), so opening these businesses may not significantly affect an epidemic outbreak.

## 6. Conclusion

The COVID-19 pandemic has resulted in the need for informed non-pharmaceutical policy interventions by public health units. We presented an agent-based modelling framework that was developed to better understand the effect of public health policy and guide their future use. The model is implemented as an object-oriented Monte Carlo simulation in Python, QUABOOM, and tracks a population of agents during an epidemic. The model implements network-like features by having households and different levels of interaction sites with which agents are associated. This allows us to model the effects of capacity restrictions on interaction sites such as workplaces, restaurants, gyms, and grocery stores. In this work, we used the code to examine capacity restrictions.

In order to define an epidemic threshold, and to compare the epidemic dynamics to real data, we introduced a definition of the basic reproduction rate, ℛ_0_, that we can compute from data and the simulation. We also introduced a new quantity, *i*_*sh*_, the number of interactions per site per hour, that allows us to develop rich interpretations of the simulations, independent of the size of the population that was used in the simulation.

We presented a new methodology for quantitatively examining capacity restrictions that would prevent an epidemic outbreak from a small number of infected agents. Resulting from the study of our model, we propose that capacity restrictions should be implemented in a targeted approach that depends on the size of the interaction site, the probability of transmission in that site, and the general attendance characteristics of that site. Our study suggests public health authorities can tailor capacity restrictions to those businesses with the largest number of interactions per hour, as we found that smaller interaction sites, with fewer customers, have little impact on the epidemics. We believe this method can be used as a new tool by public health authorities during pandemics to set appropriate restrictions based on the level of interaction sites and vaccination rates in the population. This may allow interaction sites with low risks of transmission to remain open. We believe this targeted approach is beneficial for communities and their economy, in addition to being effective at stopping the spread of an infectious virus and protecting individuals.

## Data Availability

All data produced in the present study are available upon reasonable request to the authors.

## 7. Acknowledgements

We acknowledge the support of the Department of Physics, Engineering Physics & Astronomy at Queen’s University through a research initiation grant, the Queen’s University Arts and Science Research Fund, the Queen’s University Bartlett Student Initiatives Fund, and the Natural Sciences and Engineering Research Council of Canada, funding reference number SAPIN-2017-00023.

## Footnotes

1 If someone attends a site the entire time it is open, such as a work-place, *h* = 1, if a person attends a site for 5 h when the site is open for 10 h, *h* = 2, and so on.

## Notes

### Competing Interest Statement

The authors have declared no competing interest.

